# Risk of Depression in Family Caregivers: Unintended Consequence of COVID-19

**DOI:** 10.1101/2020.06.15.20131532

**Authors:** Stephen Gallagher, Mark A. Wetherell

**Author notes:** **Corresponding Author:** Stephen Gallagher, Department of Psychology, University of Limerick, Castletroy, Co. Limerick, Republic of Ireland.

## Abstract

**Background:** COVID-19 is likely to exacerbate the symptoms of poor mental health family caregivers. To investigate whether rates of depression increased in caregivers during COVID-19 and whether the unintended consequences of health protective measures, i.e., social isolation, exacerbated this risk. Another aim was to see if caregivers accessed any online/phone psychological support during COVID.

**Method:** Data (1349 caregivers; 7527 non-caregivers) was extracted from Understanding Society, UK population level dataset. The General Health Questionnaire cut-off scores identifying those with and without depression were our primary outcome.

**Results:** After adjustment for confounding caregivers had a higher risk of having depression compared with non-caregivers, Odds ratio (OR) = 1.22 (95% confidence interval (95% CI), 1.05-1.40)), *p*=.008 evidenced by higher levels of depression pre-COVID-19 (16.7% vs 12.1%) and during COVID-19 (21.6% vs 17.9%), respectively. Further, higher levels of loneliness increased the risk of depression almost 4-fold risk in caregivers, OR = 3.85 (95% confidence interval (95% CI), 3.08-4.85)), *p*<.001), while access to therapy attenuated the risk (47%. While 60% of caregivers with depression reported not accessing any therapeutic support (e.g., online or face to face) during COVID-19.

**Conclusion:** COVID-19 has had a negative impact on family caregivers’ mental health with loneliness a significant contributor to caregiver’s depression. However, despite these detriments in mental health, the majority of caregivers do not access any online or phone psychiatric support. Reducing feelings of isolation therefore provides an opportunity for psychiatric services and health care professionals to support at-risk caregivers.

## Introduction

The COVID-19 pandemic has obvious widespread effects on physical health; however, as the pandemic continues, there is also an increasing and significant impact upon mental health. ^1^ For example, a recent-meta-analysis has found evidence of higher rates of depression in frontline workers during the pandemic relative to non-pandemic population norms. ^2^ Wide-scale public health interventions have been implemented internationally to contain the COVID-19 outbreak (e.g., school and business closures, physical distancing measures, quarantine, shielding^1^ or cocooning of at risk individuals), alongside curtailment of many health and social non-emergency services. However, these inventions are likely to have unintended consequences, especially for those who are vulnerable psychiatrically. In response, a recent consortium of psychiatrists and psychologists has strongly advocated for concerted efforts to research the impact of the pandemic on those groups that are most vulnerable. ^3^ Family caregivers, those that provide long-standing care for a family member, are an integral part of the care of the medically and psychiatrically vulnerable. The caregiver experience entails continuous demands (e.g., provision of personal, health and social care to relatives) which can be extensive both physically and emotionally, leading to a significant stress burden. ^4^ While some caregivers cope, others do not, and are at increased risk of psychological morbidity such as the development of depression ^4, 5.^

As such, family caregivers themselves have been identified as a psychiatrically vulnerable group, ^3, 5^ especially when their own needs are not met.^3^ This is an increasingly likely scenario during Covid-19 where strict limitations on movement; curtailment of non-emergency medical treatment; and changes to delivery (e.g. online or phone supports), or cessation of health and social care services is putting increasing pressure on caregivers to deliver care beyond their usual caring responsibility. ^3, 6, 7^ Caregivers routinely report having minimal opportunity to respite, little time for self-care, and increased social isolation, which together are likely to increase their risk of depression. With restrictions likely to be extended into the future, in some guise or another, this risk will increase; the impact of COVID-19 on caregivers is therefore clearly warranted. In addition, the wellbeing of the caregiver goes hand-in-hand with those for whom they provide care. If the ability to provide care and support to their family members is hindered due to their own health crisis, research has identified a greater risk of institutionalisation of loved ones, as well as additional economic, health and social costs. ^8^ It is well-documented that stress can increase vulnerability to physical disease^9,^ making caregivers at potentially greater risk of developing COVID-19. It is therefore important to identify family caregivers who are experiencing a deterioration in their mental health, and understand the factors that contribute to their increased risk (i.e., social isolation, having to provide extra care), as these may reduce their vulnerability and inform future practice and treatment.^3^

Thus, the present longitudinal case-control population-level study will explore changes in levels of depression, a common marker of carer-related psychological morbidity, and other potential contributory factors (i.e., loneliness), in family caregivers and non-caregivers assessed pre-COVID-19 and during COVID-19. We hypothesised that 1) family caregivers would have a higher rate of depression at both time points relative to non-caregivers; 2) that family caregivers who feel more isolated/lonely and who are caring more will have higher risk of being depressed. Finally, given that access to treatment may have been affected or altered (e.g., online/phone) in response to COVID-19, we also examined if caregivers with and without depression were availing of psychiatric or psychological support during COVID-19.

## Methods

### Study design and participants

A longitudinal study design was employed by using two waves of the *Understanding Society/* UK Household Longitudinal Study (UKHLS). This is a longitudinal survey of approximately 40,000 households in the United Kingdom; data were extracted from Wave 9 which was pre-COVID-19 (2017-2019)^10^ and from the specially commissioned Understanding Society COVID-19 Wave (May, 2020).^11^ All participants gave informed consent and ethics were obtained by the University of Essex, UK from National Research Ethics Service (NRES) Oxfordshire REC A (08/H0604/124). Caregiving was ascertained by asking participants: “Is there anyone in your own home who is sick, disabled or elderly whom you look after or give special help to” which had a Yes/No format. A similar question was asked for those caring outside the home. Those answering Yes, to both were pooled as one homogenous caregiver group. ^12^ For analysis, caregivers and non-caregivers had to have participated in both Waves. The final sample was N= 7527 (n=1349 caregivers). Relationship, ethnicity, and job status were dichotomized (e.g., married/partnered vs single/divorced/widowed; White vs Non-White; employed vs unemployed/retired). Similarly, living arrangements were ascertained by asking if they lived with partner (yes/no), and how many in several age brackets were living there: ages 0-4, 5-15, 16-18, 19-67, and 70 +. These were totalled and recoded into, 0= living alone, and 1 = living with others (see Table 1 for group characteristics).

**Table 1.** Sociodemographics, health and outcome variables across caregivers groups

| Variable | Non-carers<br>(N=6178) | Carers (N=1349) | Test of difference |
| --- | --- | --- | --- |
| Age | 47.5 (17.70) | 52.8(14.79) | F (1,9768) = 142.56, $p < .001$ |
| Married/Partnered % | 74.2 | 79.5 | $\chi^2 (1) = 16.82, p < .001$ |
| Sex (female) % | 51.3 | 61.5 | $\chi^2 (1) = 21.00, p < .001$ |
| Ethnicity % (White) | 92.6 | 93.6 | $\chi^2 (1) = 2.10, p = .14$ |
| Income (Monthly £) | 1,700 (1,785) | 1,635 (1,585) | F (1,9768) = 2.03, $p = .15$ |
| Employed % (no) | 35.9 | 5.2 | $\chi^2 (1) = 50.89, p < .001$ |
| Disability % (yes) | 31.2 | 43.9 | $\chi^2 (1) = 100.38, p < .001$ |
| Living alone % (yes) | 2.0 | 3.4 | $\chi^2 (1) = 100.38, p < .001$ |
| PreCOVID-19 Lonely (often) | 7.5 | 8.0 | $\chi^2 (2) = 11.90, p < .001$ |
| COVID-19 Lonely (often) | 7.1 | 8.2 | $\chi^2 (2) = 2.73, p = .25$ |
| Therapy access % (no) | 42.1 | 59.7 | $\chi^2 (2) = 375.5, p < .001$ |
| PreCOVID-19 Depression % (yes) | 12.1 | 16.7 | $\chi^2 (1) = 26.68, p < .001$ |
| COVID-19 Depression % (yes) | 17.9 | 21.6 | $\chi^2 (1) = 12.45, p < .001$ |

### Outcomes

Depression was our primary outcome and captured in both Waves by the 12-item General Health Questionnaire (GHQ).^13^ Items (e.g., unhappy or depressed) are scored as 1= not at all; 2= no more than usual; 3= rather more than usual; 4= much more than usual. Responses of 1 or 2 are scored as 0, and responses of 3 or 4 are scored as 1. A total score ≥6 is specific and sensitive at identifying those with or without a depressive disorder. ^14^

### Access to therapy during COVID-19

Access to therapy was a secondary aim and was assessed by asking: “In the last month, have you access counselling or talking therapy?”, response categories were: 1= Yes, in person; 2 = Yes, by telephone or online; 3 = Yes, group sessions; 4 = No; and 5 = Not required.

### Loneliness

Loneliness at both time-points was assessed by a single item: “In the last 4 weeks, how often did you feel lonely?” with three responses, 1= Hardly ever or never; 2= Sometimes; 3= Often.

### Extra Caring

More caring responsibility during COVID-19 was assessed by the question “How has the help and support you receive from family, friends or neighbours who do not live in the same house/flat as you changed?; responses were: 1 = There has been no change; 2 = I receive more help from some people who previously helped me; 3 = I receive less help from some people who previously helped me; 4 = I currently receive help from family, friends or neighbours who did not previously help me;5 = Other.

### Statistical Analysis

Weights were calculated to provide a representative national sample, taking into account survey design and non-response. For more information on the weighting system, see the main Understanding Society COVID-19 report.^**11**^ No outliers were observed and data was normally distributed. There was a low response rate (n= 2) for the access to therapy response face to face therapy’ (1 depressed and 1 not depressed) and n =1 (depressed) for group sessions. Thus, we pooled these with the online/phone group response as all respondents had access to therapy, to produce access to therapy three ‘access to therapy’ categories: 1= accessed online or phone treatment; 2= no access; and 3= did not require treatment. Tests of differences were used to examine group differences on sociodemographic, health and outcome variables. Logistic regression was used to examine the predictors of risk of depression in caregivers relative to non-caregivers. In this analysis, non-caregivers and those without depression were both dummy coded as 0, while caregivers and those with depression were coded at 1. Confounding variables (i.e., health and sociodemographics, pre-existing depression and loneliness) were entered in step 1 of the model and caregiver groups (non-caregivers vs caregivers) entered at Step 2. This was followed by another logistic regression on caregivers only to examine the predictors of depression risk with the same confounding factors entered in Step 1, pre-COVID depression, and loneliness at Step 2, and current COVID-19 related loneliness, access to treatment and more or less caring during the crisis in Step 3. Odds ratio (OR) is the effect size.

## Results

As can be seen in Table 1, caregivers were slightly older, more likely to be married/partnered, be female, be unemployed/retired during COVID-19, live alone, and have a heath condition/disability in comparison to non-caregivers. As expected, rates of depression increased during the pandemic for both groups. However, caregivers had higher rates of depression (16.7%, 21.6%), than non-carers (12.1%, 17.9%), at pre-COVID-19 and during COVID-19 respectively.

In logistic regression, after controlling for confounding factors (See Table 1 for group differences) in Step 1, caregivers had a 21% greater risk of being depressed compared to non-caregivers; OR = 1.22 (95% confidence interval (95% CI), 1.05-1.40)), *p* =.008. In within caregiver group analysis (See Table 2), after controlling for confounding variable in Step 1, and Step 2, we found that the key predictor of this excess risk was current loneliness such that those who felt lonelier during the current crisis had an almost 4-fold risk of depression, OR = 3.85 (95% confidence interval (95% CI), 3.08-4.85)), *p*<.001. As can be seen in Figure 1, almost 80% of those who reported being lonely ‘quite often’ were depressed, whereas 90% caregivers who were not depressed reported never being lonely. Access to psychological supports was also reduced depressive risk by 43% (see Table 2). Finally, we explored how access to treatment varied across caregivers with and without depression. As can be seen in Figure 2, 60% of carers with depression said they did not access any psychological supports, while 20% of caregivers with depression reported ‘no need for supports’(χ^2^ (2) = 78.63, *p* <.001)

**Table 2.** Hierarchical logistic regression sociodemographic, health, loneliness, caring more/less and access to therapy predicting family caregiver depression.

| Variables | B | OR | *p* | 95%CI Lower | 95%CI Upper |
| --- | --- | --- | --- | --- | --- |
| **Step 1** |  |  |  |  |  |
| Age | -.01 | 0.99 | .23 | 0.97 | 1.06 |
| Partnered | -.01 | 0.70 | .94 | .61 | 1.58 |
| Gender | .84 | 2.32 | **.001** | 1.54 | 3.49 |
| Ethnicity | .77 | 2.26 | **.033** | 1.06 | 4.29 |
| Income | .00 | 1.00 | .12 | 1.00 | 1.00 |
| Employment | -.01 | 0.99 | .91 | 0.98 | 1.03 |
| Living alone | .62 | 1.85 | .11 | 0.86 | 4.00 |
| Disability | -.29 | 0.75 | .14 | 0.51 | 1.07 |
| **Step 2** |  |  |  |  |  |
| Pre COVID-Depression | 1.05 | 2.76 | **.001** | 1.83 | 4.15 |
| Pre COVID-Loneliness | .05 | 1.09 | .53 | 0.82 | 1.46 |
| **Step 3** |  |  |  |  |  |
| Access to therapy | -.56 | .57 | **.001** | 0.40 | 0.80 |
| More or less caring | .09 | 1.09 | .68 | 0.70 | .168 |
| Covid-19 loneliness | 1.35 | 3.87 | **.000** | 3.08 | 4.85 |

**Figure 1.**
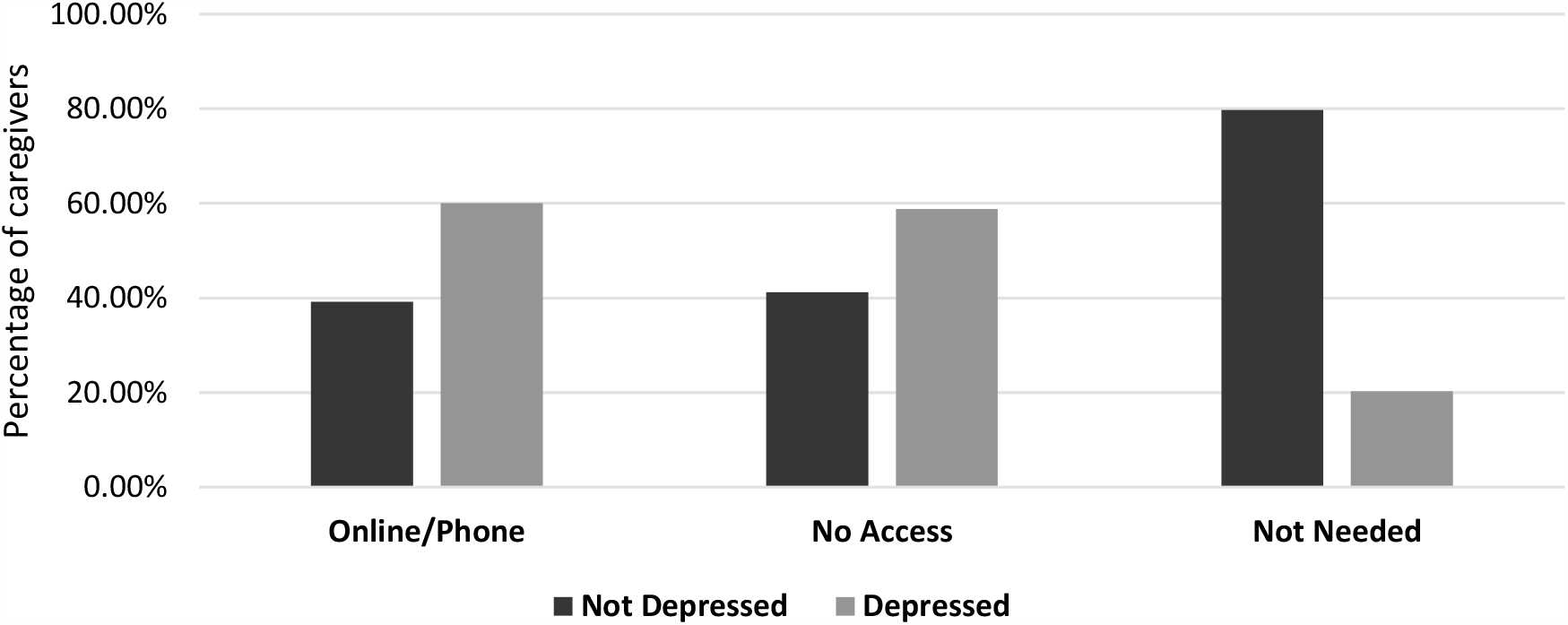
Depression and access to psychological therapy during COVID-19 in caregivers.

**Figure 2.**
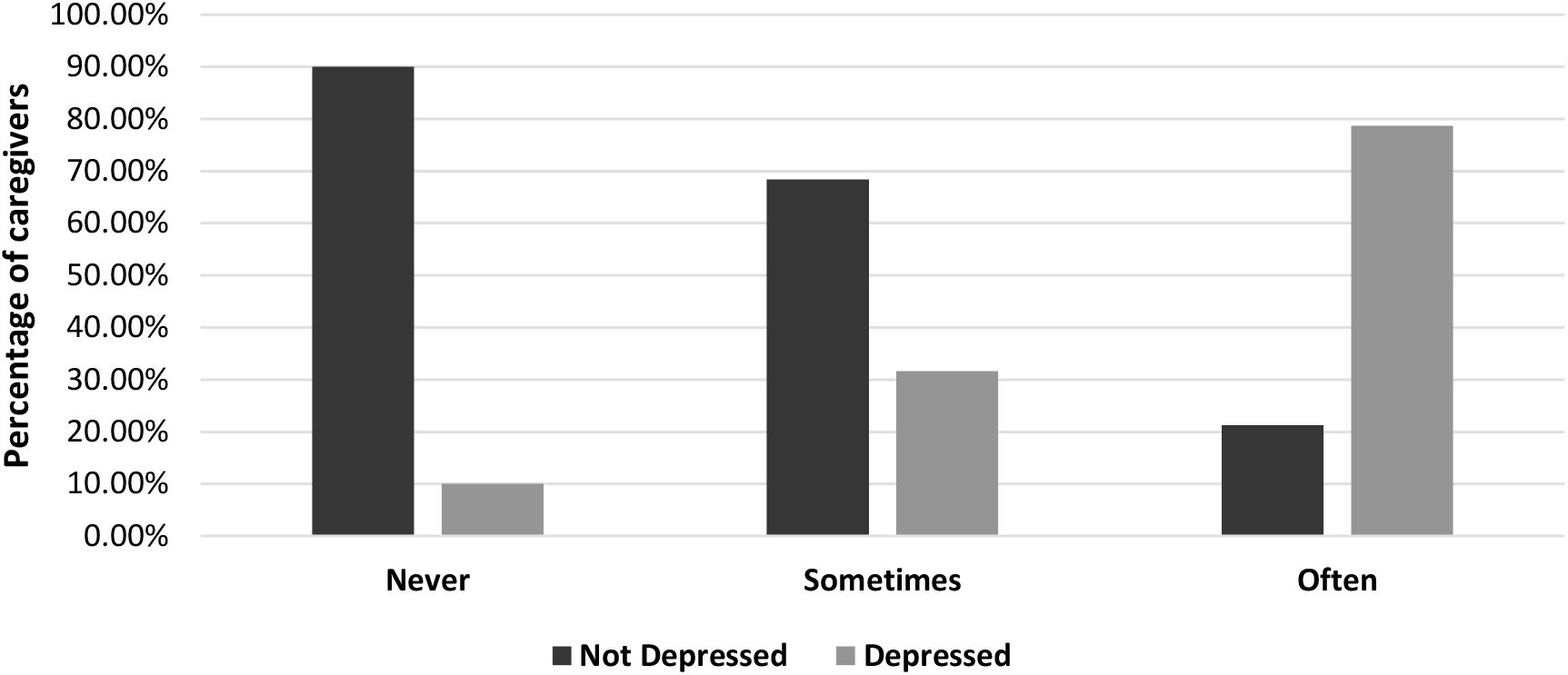
Depression and feelings of loneliness during COVID-19 in caregivers.

## Discussion

The present study demonstrates that levels of depression have increased in a large sample of UK citizens during COVID-19. Additionally, and in line with previous research, caregivers report greater levels of depression compared with non-caregivers and this is evident both pre and during COVID-19. Furthermore, current levels of loneliness were a significant predictor of depression risk while access to psychological supports attenuated the risk. However, it is worth noting that a large proportion of caregivers with depression felt they did not need psychotherapy supports.

It is likely that the public health restrictions implemented to protect people from COVID-19 are having unintended consequences, ^3, 7^ as evidenced by the fact that current feelings of loneliness and not previous loneliness, exacerbated caregivers’ depression risk. Moreover, this was evident irrespective it caregivers lived alone or not. Further, our findings also showed that while 20% of caregivers who were categorised with depression felt they did not need access to therapy, a large percentage (60%) of caregivers with depression reported having no psychological supports during the pandemic.

While our findings align with recent observations showing that risk of depression has increased during the current crisis.^2^ In the present study, we were able to demonstrate that this risk is exacerbated in family caregivers who are already mentally vulnerable, by comparing levels of depression from pre-COVID-19 to levels experienced during the current crisis. Levels of depression as indexed using a ≥6 on the GHQ ^14^ to identity those with depression, increase by 5% in family caregivers during this period. Thus, the recent call to investigate the negative effects of COVID-19 on those with existing psychiatric vulnerabilities was warranted and confirmed by the findings of this study. ^3^ More importantly, we have identified the effects of isolation, a likely consequence of restrictions aimed at reducing spread of COVID-19, as a potential contributor to increased depression. In fact, it was loneliness experienced during the pandemic and not previous levels of loneliness that was a significant contributor, with caregivers who experienced greater levels of loneliness having an almost 4-fold increased risk of depression. Further, caregivers did not report significant changes to their caring duties, suggesting that it is the unintended psychological consequences of COVID-19 restrictions, and not the physical demands of having to care more, that are contributing to levels of depression. Despite increases in levels of depression during COVID-19, and the benefits of accessing therapeutic treatments, a significant proportion of caregivers reported no access to psychological supports during this period. This suggests that for many psychologically vulnerable individuals, support is either not available or not being sought.

### Limitations

This study has several strengths, notably a longitudinal design and the use of a validated tool for the assessment of depression in population research. In response to recent calls, it also assesses an at risk group and identifies potential contributory factors linked to depression. However, these findings should be considered in light of some limitations. First, we do not have the details of the type of care-recipients’ illness/disability type which may confer an additional risk and differences in the caregiver experience and associated care burden (e.g. Alzheimers and Cancer). Second, there may be other unmeasured variables that may also explain risk of depression (e.g. worry and anxiety), and as can be seen in Figure 2, some 40% of caregivers were not categorised as depressed but were availing of treatments, and as such may have had another mental health condition. Third, while caregiving was predictive of depression at both time-points, we cannot infer causality. Increased psychiatric symptomatology and rates of common mental disorder in caregivers could also reflect shared biological vulnerabilities with their care-recipient relatives. While plausible, there is convincing evidence that it is the caregiver role itself that drives increases in psychological morbidity such as depression, ^5^ Finally, depression status was derived through a self-report scale rather than psychiatric interview. Nonetheless, we used a widely used scale, which has intrinsic value as an indicator of psychological distress, particularly in population studies such as this,^15^ but also given the central role of subjective carer appraisals in cognitive models of caregiver burden.^16^ Relatedly, a lack of formal diagnosis of depression may explain why a large proportion of the caregivers were not accessing any psychological support resources. That is, despite reporting high levels of depression using a questionnaire, they may not have identified the need for accessing support that would typically be made available following a formal diagnosis. Moreover, we do not know to whether individuals were accessing support services before COVID-19.

### Implications

Notwithstanding these limitations, this study demonstrates that caregivers are at increased risk of depression, and that feelings of isolation may increase this risk. Current and future restrictions related to social distancing will exacerbate this issue, and as such, caregivers are an especially vulnerable group during COVID-19. Given the well-established consequences of caregiving stress on physical health,^4^ caregivers may be at increased risk of contracting COVID-19, which would have detrimental effects on their ability to provide care and implications for institutionalisation of relatives as well as additional health and social care costs. The identification of caregivers as an at-risk group can therefore act as a prompt for psychiatric and mental health services to better understand the causes and consequences of psychological morbidity in this group. For example, this study has identified the potential role of perceived isolation as a risk factor for depression. As the pandemic continues caregivers may become greater users of mental health services, and the identification of risk factors may provide targets for intervention and treatment. Of significant note; however, is the observation that many of the caregivers in this study did not access any support services, despite their levels of depression. These particular findings have implications for current practice, and how to tailor or address the needs of this vulnerable group and ensure that appropriate support services are accessible.

Attempts to reduce the burden of caregiving has clear implications for the psychological and physical health of the carer, the wellbeing of the care recipient, and potential societal and financial costs. This is not a new challenge and there are ongoing endeavours to address this need. However, given the potential role of feelings of isolation as a risk factor, the ongoing impact of COVID-19 raises specific issues for caregivers and thus, efforts to alleviate this risk are needed.^17^

## Data Availability

Data Availability: The data that support the findings of this study are openly available in Understanding Society: COVID-19 Study, 2020 at http://doi.org/10.5255/UKDA-SN-8644-1 and Wave 9 at http://doi.org/10.5255/UKDA-SN-6669-11

https://ukdataservice.ac.uk/

## Authors Contribution

Professor Gallagher designed the study, accessed, screened and analysed the data. Professor Gallagher and Professor Wetherell wrote and drafted the manuscript.

## Data Availability

The data that support the findings of this study are openly available in Understanding Society: COVID-19 Study, 2020 at http://doi.org/10.5255/UKDA-SN-8644-1 and Wave 9 at http://doi.org/10.5255/UKDA-SN-6669-11

## Footnotes

1 Sheilding/Cocooning: Are concepts used in the UK and Ireland to describe social isolation procedure instructions to protect the medically vulnerable COVID-19

## References

1. Kesner L, Horacek J. Three challenges that the COVID-19 pandemic represents for psychiatry. Br J Psychiatry. 2020:1–2.

2. Pappa, S., et al., Prevalence of depression, anxiety, and insomnia among healthcare workers during the COVID-19 pandemic: A systematic review and meta-analysis. Brain Behav Immun, 2020.

3. Holmes, E.A., et al., Multidisciplinary research priorities for the COVID-19 pandemic: a call for action for mental health science. Lancet Psychiatry, 2020.

4. Lovell, B. and M.A. Wetherell, The cost of caregiving: endocrine and immune implications in elderly and non elderly caregivers. Neurosci Biobehav Rev, 2011. 35(6): p. 1342–52.

5. Smith, L., et al., Mental and physical illness in caregivers: results from an English national survey sample. Br J Psychiatry, 2014. 205(3): p. 197–203.

6. Bowers, B., K. Pollock, and S. Barclay, Administration of end-of-life drugs by family caregivers during covid-19 pandemic. BMJ, 2020. 369: p. m1615.

7. Phillips, D., et al., The invisible workforce during the COVID-19 pandemic: Family carers at the frontline [version 1; peer review: awaiting peer review]. HRB Open Research, 2020. 3(24).

8. Reinhard, S.C., et al., Supporting Family Caregivers in Providing Care, in Patient Safety and Quality: An Evidence-Based Handbook for Nurses, R.G. Hughes, Editor. 2008: Rockville (MD).

9. McEwen, B.S., Stress, adaptation, and disease. Allostasis and allostatic load. Ann N Y Acad Sci, 1998. 840: p. 33–44.

10. University of Essex, Institute for Social and Economic Research, and K.P. NatCen Social Research, University of Essex, Institute for Social and Economic Research, NatCen Social Research, Kantar Public. (2019). Understanding Society: Waves 1-9, 2009-2018 and Harmonised BHPS: Waves 1-18, 1991-2009. [data collection]. 12th Edition. UK Data Service. SN: 6614, http://doi.org/10.5255/UKDA-SN-6614-13. 2019.

11. University of Essex and I.f.S.a.E. Research., Understanding Society: COVID-19 Study, 2020. [data collection]. UK Data Service. SN: 8644, http://doi.org/10.5255/UKDA-SN-8644-1. 2020

12. Gallagher, S., A.C. Phillips, and D. Carroll, Parental stress is associated with poor sleep quality in parents caring for children with developmental disabilities. J Pediatr Psychol, 2010. 35(7): p. 728–37.

13. Goldberg, D.P., et al., The validity of two versions of the GHQ in the WHO study of mental illness in general health care. Psychol Med, 1997. 27(1): p. 191–7.

14. Lundin, A., et al., Validity of the 12-item version of the General Health Questionnaire in detecting depression in the general population. Public Health, 2016. 136: p. 66–74.

15. Pierce, M., et al., Says who? The significance of sampling in mental health surveys during COVID-19. The LANCET Psychiatry 2020. June https://doi.org/10.1016/S2215-0366(20)30237-6

16. Szmukler, G.I., et al., Caring for relatives with serious mental illness: the development of the Experience of Caregiving Inventory. Soc Psychiatry Psychiatr Epidemiol, 1996. 31(3-4): p. 137–48.

17. Luykx, J.J., C.H. Vinkers, and J.K. Tijdink, Psychiatry in Times of the Coronavirus Disease 2019 (COVID-19) Pandemic: An Imperative for Psychiatrists to Act Now. JAMA Psychiatry, 2020.

